# Utilisation Patterns of Cardiovascular Medicines Among Outpatients at a Tertiary Cardiac Institute in Tanzania (2017-2022)

**DOI:** 10.64898/2025.11.30.25341295

**Authors:** Justine L. Vasco, Raphael Z. Sangeda, Tusaligwe Mbilinyi, Naizihijwa Majani, Reuben Kato Mutagaywa, Peter Richard Kisenge

## Abstract

**Background:** Cardiovascular diseases are among the leading causes of morbidity and mortality worldwide. Routine quantification of cardiovascular medicine utilisation is essential for stewardship, formulary decision-making and procurement planning.

**Objective:** To describe outpatient utilisation of cardiovascular medicines at the Jakaya Kikwete Cardiac Institute (JKCI) using ATC/DDD methodology.

**Methods:** This was a retrospective longitudinal analysis of outpatient prescriptions (2017/2018-2021/2022). Medicines were classified according to the ATC system and quantified as Defined Daily Doses (DDD) per 100 outpatient days. The analyses progressed hierarchically from ATC levels 1 to 5. DU metrics (DU50/DU90) were used to assess the concentration. Monthly utilisation was examined using Seasonal Trend decomposition using Loess (STL). Statistical analyses were performed using SPSS (version 26).

**Results:** Across the study period, the cumulative utilisation of cardiovascular medicines totalled 360.55 DDD per 100 outpatient days. Annual intensity peaked in 2018/2019 (82.92 DDD per 100 outpatient-days), was the second-highest in 2020/2021 (82.39), and was lowest in 2017/2018 (55.68). At ATC level 5, amlodipine (C08CA01), candesartan (C09CA06) and clonidine (C02AC01) were the principal contributors. The DU50 segment comprised five molecules (amlodipine, candesartan, clonidine, furosemide, and atorvastatin), whereas DU90 ranked 18th, indicating concentrated utilisation within a small number of agents. The STL showed a gradual secular rise with no statistically significant month-specific seasonality.

**Conclusion:** Outpatient cardiovascular medicine utilisation at JKCI was concentrated within a narrow set of anti-hypertensive agents, with a cumulative burden of 360.55 DDD per 100 outpatient-days and a fiscal-year peak in 2018/2019. Institutionalising routine outpatient DDD surveillance may support stewardship, rational prescribing, and more equitable access to high-impact cardiovascular medicines in Tanzania.

## 1.0 Introduction

Cardiovascular diseases (CVDs) are the leading contributors to morbidity and mortality worldwide, including in sub-Saharan Africa. Despite the expansion of specialised cardiac services, evidence from low- and middle-income countries (LMICs) continues to demonstrate persistent inequities in the availability, affordability, and rational use of cardiovascular medicines [1–3]. Systematic reviews indicate that most LMICs fail to meet the WHO benchmark of 80% availability for essential cardiovascular drugs, and affordability often requires multiple days’ wages for a one-month supply [1,2].

Outpatient services are the principal point for long-term management of CVDs, yet most pharmacoepidemiological studies have concentrated on inpatient or intensive care settings. Emerging evidence from Asia, Africa, and the Middle East has revealed variable adherence to anti-hypertensive and lipid-lowering guidelines, frequent polypharmacy among older adults, and regional disparities in the uptake of evidence-based therapies [4–7]. Even in multi country analyses, secondary prevention medicines remain underused across all income strata, highlighting the systemic barriers to access and prescribing [8–11].

Drug utilisation research, based on the World Health Organisation Anatomical Therapeutic Chemical (ATC) and Defined Daily Dose (DDD) methodology, provides a standardised means of quantifying and comparing medicine use across healthcare settings. While DDD per 1,000 inhabitants per day is the most common measure for population level analysis, outpatient hospital utilisation is better captured using DDD per 100 outpatients per day, which reflects actual service utilisation rather than population exposure [12,13]. This metric differs from the DDD per 100 bed days used for inpatient surveillance and is particularly suited for longitudinal pharmacoepidemiological assessments in ambulatory care settings [14].

In Tanzania, the increasing burden of cardiovascular disease has prompted the development of tertiary cardiac care capacity through institutions such as the Jakaya Kikwete Cardiac Institute (JKCI). However, systematic evidence on the use of outpatient cardiovascular medicine remains limited. Therefore, this study quantified the utilisation of cardiovascular drugs among outpatients at JKCI between 2017 and 2022 using DDD per 100 outpatients per day. These findings align with global LMIC trends in access, affordability and secondary prevention, inform rational medicine use and policy interventions.

## 2.0 Methodology

### 2.1 Study design

This was a retrospective longitudinal pharmacoepidemiologic study assessing the utilisation of cardiovascular medicines among outpatients attending a tertiary cardiac care facility in Tanzania from July 2017 to June 2022.

### 2.2 Study setting

The study was conducted at the Jakaya Kikwete Cardiac Institute (JKCI), a specialised tertiary hospital located in Upanga West, Dar es Salaam, Tanzania. The JKCI provides diagnosis, treatment, and follow-up care for a wide range of cardiovascular diseases (CVDs). The institute operates an electronic health information system (MedPro) that captures detailed patient-level prescription data. Data.

### 2.3 Study population

The study population comprised all outpatient encounters in which ≥1 cardiovascular medicine (Anatomical Therapeutic Chemical [ATC] class C) was dispensed during the study period. Encounters were classified as outpatient visits if no inpatient bed days were recorded for that encounter.

### 2.4 Data source

Patient-level dispensing records were extracted from the MedPro hospital information system at the JKCI. The extracted fields included patient ID, visit number, age, sex, visit date, ATC generic name, dose form, strength, pack size, quantity dispensed, route of administration, and mode of payment (sponsor category). The data were anonymised before analysis.

### 2.5 Exclusion criteria

Topical medicines (creams, ointments, lotions, shampoos and other dermatological formulations) were excluded. Oral, parenteral and other systemic formulations were retained.

### 2.6 Data collection, cleaning and analysis

Data were extracted from the JKCI MedPro database in collaboration with the Institute’s ICT personnel. The dataset included demographic and prescription information for all outpatients between 2017 and 2022. Key variables included age, sex, diagnosis, and medication details (generic name, route of administration, dosage form, strength, pack size and quantity dispensed). Medicines were classified using the Anatomical Therapeutic Chemical (ATC) system, and utilisation was quantified using the Defined Daily Dose (DDD) methodology, as recommended by the WHO Collaborating Centre for Drug Statistics Methodology. Topical preparations (e.g. lotions, creams, ointments and shampoos) were excluded [12,13].

Drug utilisation analyses followed the ATC hierarchy. Utilisation was first summarised at ATC Level 1 and then progressively disaggregated through Levels 2, 3, and 5, where specific chemical substances were defined. This hierarchical drill-down is standard in DU/DDD pharmacoepidemiology to separate system-level burden, therapeutic grouping and molecule-level utilisation. Operationally, the hierarchy moves from (i) system class (Level 1, for example, C = cardiovascular), (ii) therapeutic main groups (Level 2), (iii) pharmacological/chemical sub-classes (Levels 3–4), and (iv) individual chemical substances (Level 5), which are the basis for the DU50/DU90 assessment. Focused antibiotic DDD analyses have also been performed in the same population [14].

Outpatient utilisation intensity was expressed as DDD per 100 outpatient days. This denominator was anchored to the actual number of cardiovascular outpatient encounters at the JKCI and was calculated using the following formula:

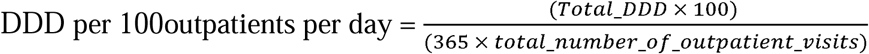

This allows for a standardised comparison of utilisation across fiscal years and patient subgroups within the facility. Subgroup descriptive summaries were pre-specified for the sponsor category, sex, age group, fiscal year, and episode persistence. For each subgroup, the crude mean DDD per 100 outpatient days and cumulative contributions were computed. These comparisons were descriptive and unadjusted for confounding; their purpose was to characterise the empirical distribution before the inferential modelling.

Time-series analysis of monthly utilisation (July 2017 – June 2022) employed Seasonal–Trend decomposition using Loess (STL) to characterise long-range trends. Month-specific seasonal effects were assessed using a one-way ANOVA across calendar months.

Data transformation was performed in Microsoft Power BI (Power Query), and statistical analyses, including ANOVA, STL-based trend estimation, curve estimation, and forecasting, were performed using IBM SPSS Statistics version 26.

### 2.7 Ethical consideration

Ethical approval was granted by the Muhimbili University of Health and Allied Sciences (MUHAS) Institutional Review Board (Reference DA.25/111/01B/184; February 2022). JKCI granted permission to access anonymised data. No personal identifiers were retained in the analytical dataset. The requirement for patient consent was waived because this study used fully anonymised routine care data.

## 3.0 Results

### 3.1 Descriptive characteristics and crude comparisons

A total of 52,199 outpatient cardiovascular medicine encounters were analysed; 52,178 (99.96%) had non-missing DDD per 100 outpatient-day estimates. Females contributed 28,604 (54.8%) encounters, whereas males contributed 23,595 (45.2%). The age distribution was dominated by children aged 0–9 years (13,689; 26.2%), followed by adults aged 60–69 years (9,613; 18.4%) and 50–59 years (8,936; 17.1%). More than half of the encounters (29,699; 56.9%) had “one-year only” attendance patterns (initiated and completed within the same fiscal year), whereas 22,500 (43.1%) continued into subsequent years. The NHIF category represented the largest sponsor group (30,500, 58.4%), followed by private cover (19,107, 36.6%).

Crude mean utilisation differed significantly across sponsor groups (ANOVA, p < 0.001), with the highest DDD per 100 outpatient days observed among NHIF beneficiaries and the lowest among the private and other categories (Table 1). Females had slightly higher mean values than males (0.00712 vs. 0.00665; p = 0.012). A clear age gradient was observed (p < 0.001), with the lowest mean in children aged 0–9 years and the highest in adults aged 60–69 years.

**Table 1:**
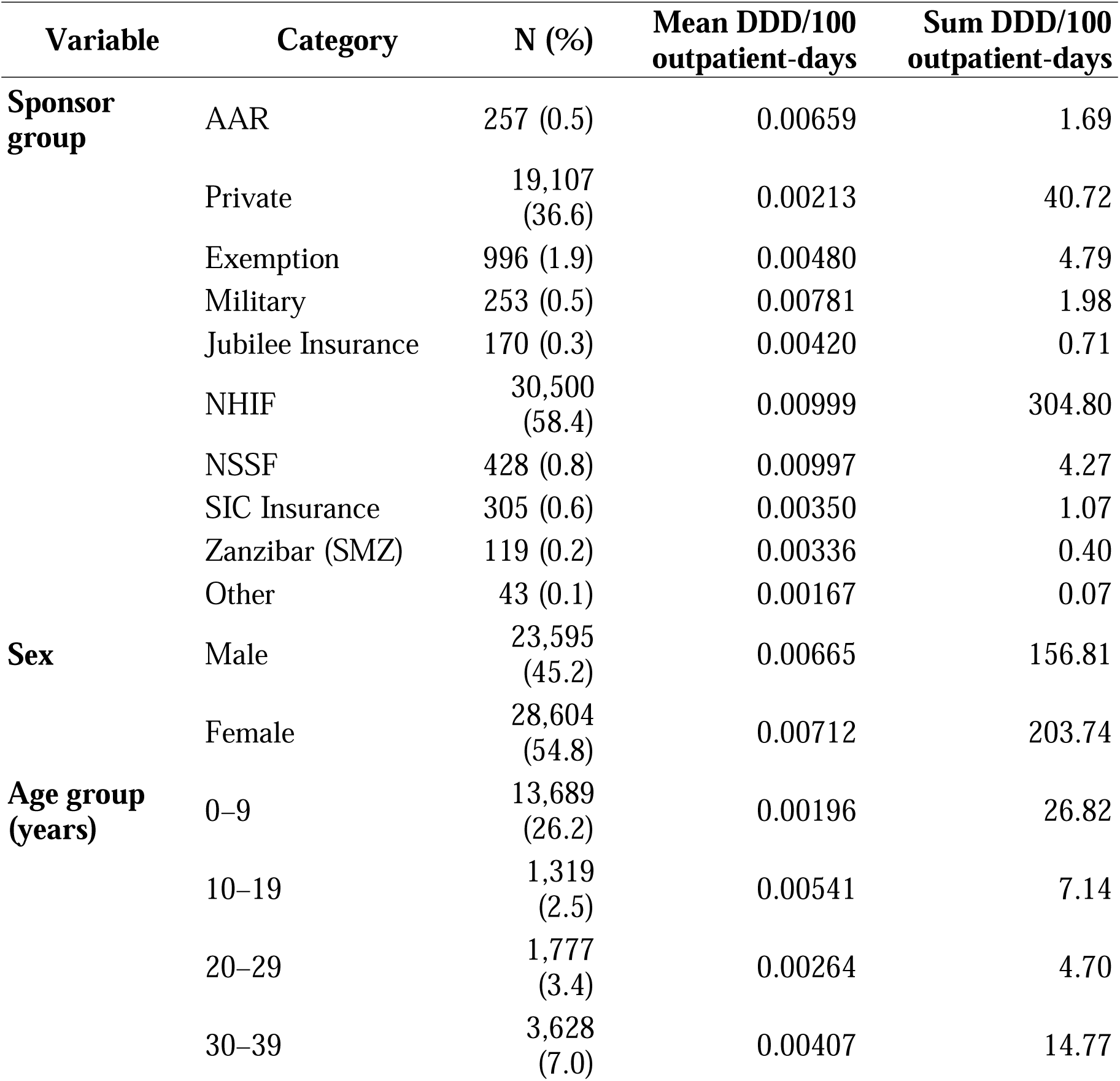
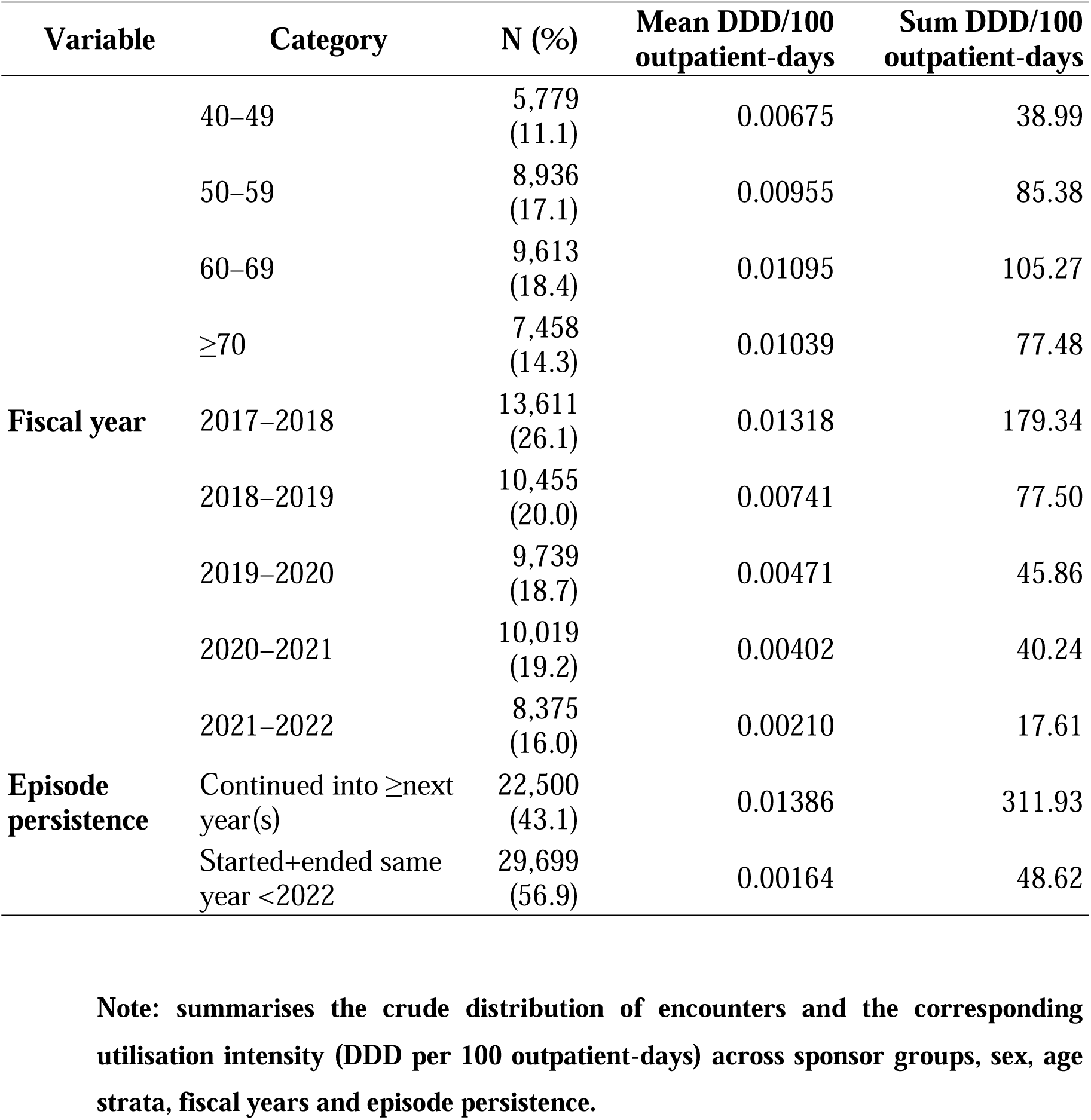
Characteristics of outpatients who received ≥1 cardiovascular medicine (ATC Class C) and utilisation intensity (DDD per 100 outpatient-days) across subgroups at the Jakaya Kikwete Cardiac Institute (JKCI), FY 2017/2018–2021/2022 ( N = 52,199).

Utilisation intensity also declined across fiscal years, from 0.01318 DDD per 100 outpatient-days in FY 2017/2018 to 0.00210 in FY 2021/2022 (ANOVA p < 0.001) (Table 1).

A total of 1,119,041 outpatient prescription records **were** analysed across all ATC level 1 therapeutic classes (A–V) during the five-year study period, reflecting the full spectrum of medicines dispensed to patients attending JKCI outpatient clinics. These records included agents for the alimentary tract and metabolism (A), anti-infectives (J), cardiovascular medicines (C), nervous system agents (N), and several other therapeutic categories, represented in smaller proportions.

Across all ATC level 1 categories, cardiovascular medicines (ATC class C) were consistently the most utilised group throughout the review period, followed by anti-infectives (J) and agents for the alimentary tract and metabolism (A) (Figure 1). The rankings of the major ATC level 1 classes remained stable and did not change significantly across fiscal years.

**Figure 1:**
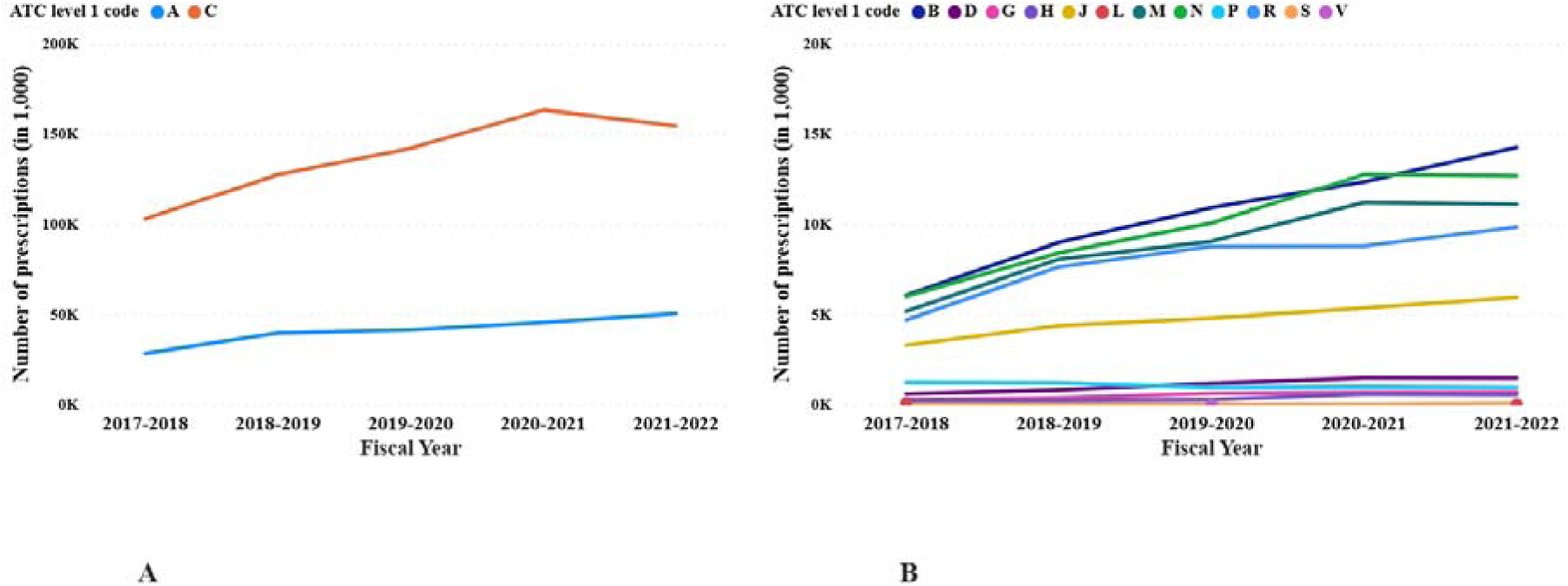
Number of prescriptions for ATC level 1 medicine classes used at JKCI, fiscal years 2017/2018–2021/2022. Key for ATC codes level 1: A = Alimentary Tract and Metabolism; B = Blood and Blood Forming Organs; C = Cardiovascular System; D = Dermatologicals; G = Genito-urinary System and Sex Hormones; H = Systemic Hormonal Preparations Excluding. Sex hormones and insulin; J = anti-infectives for systemic use; L = antineoplastic and immunomodulating agents; M = musculoskeletal system; N = nervous system; P = antiparasitic products, insecticides, and repellents; R = respiratory system; S = sensory organs; and V = various.

Across fiscal years, the total number of outpatient prescriptions increased from 158,401 in 2017–2018 to a peak of 262,499 in 2020–2021, before stabilising at 261,875 in 2021–2022. Cardiovascular medicines (ATC class C) followed a similar pattern, rising from 102,739 in 2017–2018 to 163,027 in 2020–2021, and then declining slightly to 154,241 in 2021–2022 (Figure 2).

**Figure 2:**
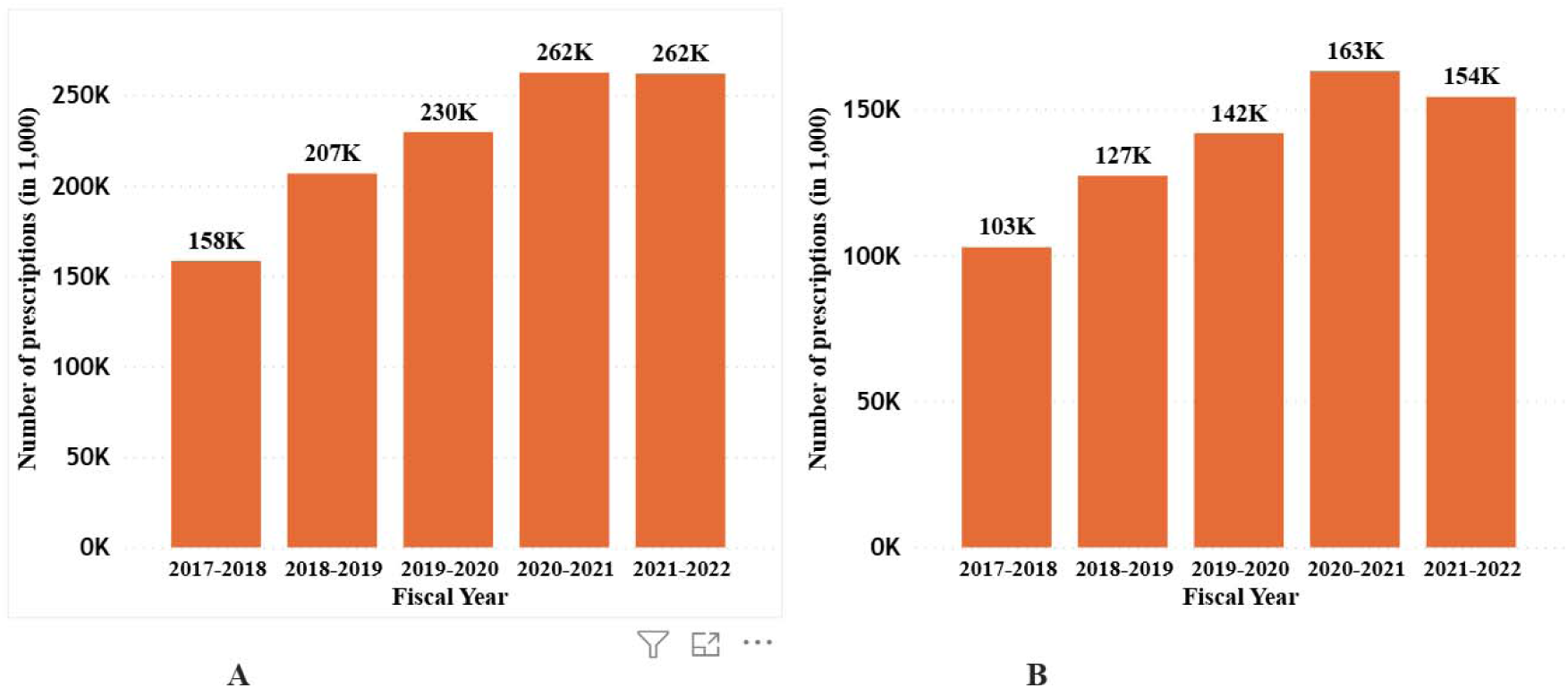
Annual outpatient utilisation counts at JKCI by fiscal year (2017/2018–2021/2022). Panel A: Total prescriptions (all ATC classes). Panel B: Prescriptions of cardiovascular medicines (ATC class C only). The values represent the total number of annual prescription rows extracted from the patient pharmacy records.

The full distribution of all ATC level 5 medicines prescribed during the study period (265 items in total), ranked by frequency of use, depicts that the five most frequently dispensed medicines were amlodipine (C08CA01), candesartan (C09CA06), furosemide (C03CA01), spironolactone (C03DA01), and bisoprolol (C07AB07), which together accounted for approximately 29 percent of all prescriptions (Supplementary Table 1).

At the annual level, cardiovascular utilisation was summarised using two complementary indicators: (i) cumulative annual DDD per 100 outpatient-days and (ii) number of unique outpatients (UHID-based) who received ≥1 cardiovascular medicine during that year. Starting from 2017/2018, both indicators rose to a peak in 2018/2019 (84.95 DDD per 100 outpatient-days; 38,468 unique outpatients), declined thereafter, and reached the lowest values again toward the start of the series (56.54 DDD per 100 outpatient-days; 28,298 unique outpatients in 2017/2018) (Figure 3).

**Figure 3:**
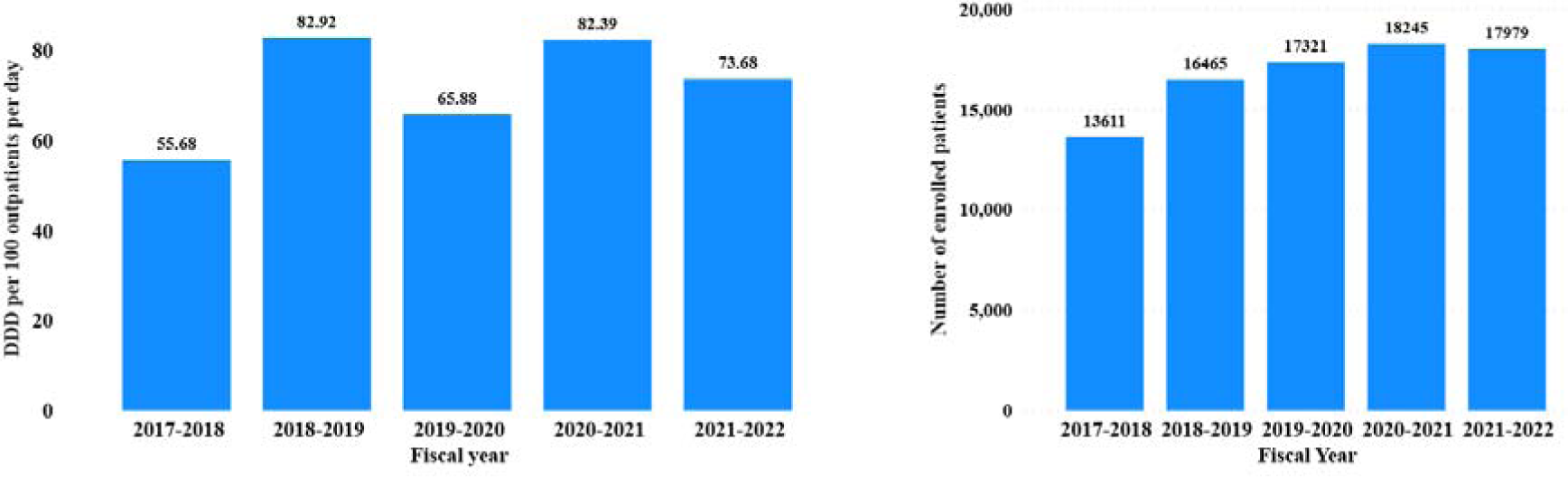
Annual cardiovascular outpatient utilisation at JKCI from 2017/2018 to 2021/2022. Panel A: Cumulative utilisation intensity expressed as Defined Daily Doses per 100 outpatient days (facility-level denominator). Panel B: Number of unique annual outpatient cardiovascular patients, defined by Unique Hospital Identification (UHID; one record per patient per fiscal year). Note: Table 1 uses encounter-level prescription rows; therefore, the denominators differ from the UHID-based patient counts used in Panel B. per 100 outpatient days.

At ATC Level 2, consumption intensity varied across fiscal years, with the highest cumulative utilisation observed for agents acting on the renin–angiotensin system (C09; 114.75 DDD per 100 outpatient-days), followed by calcium channel blockers (C08; 65.71) and diuretics (C03; 52.60) (Figure 4).

**Figure 4:**
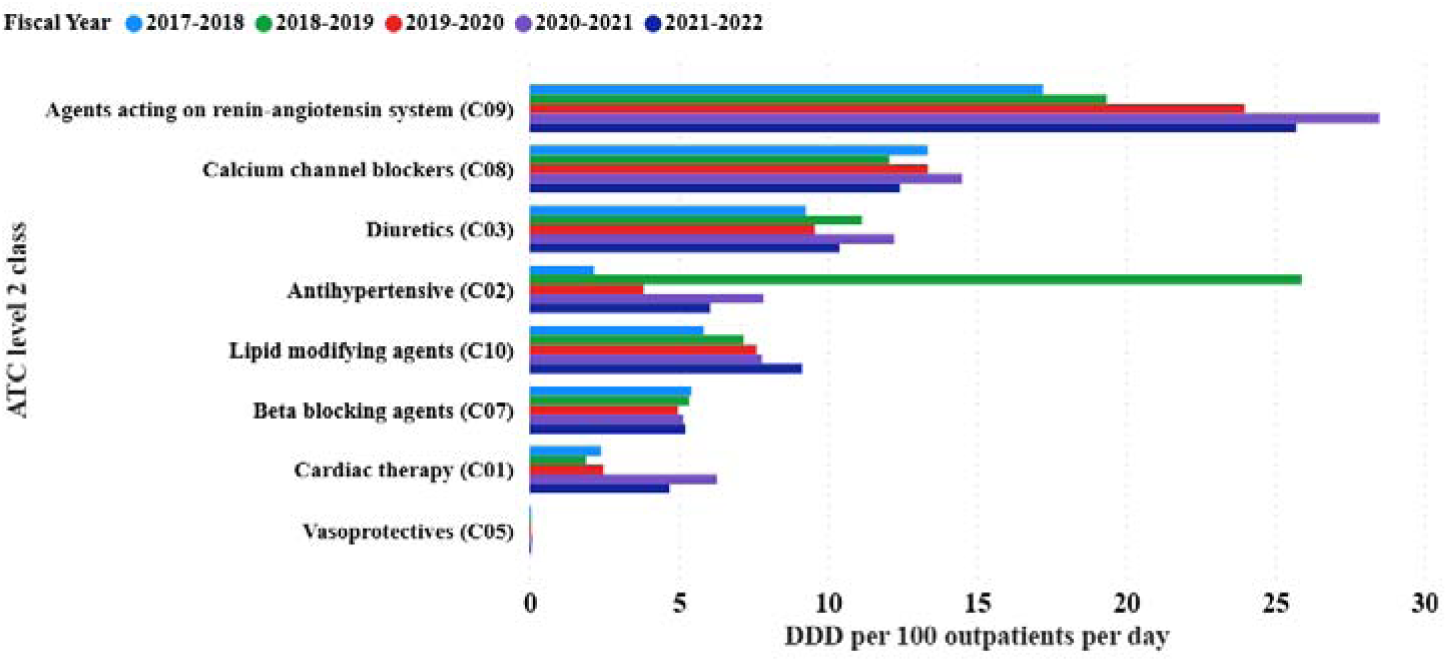
Annual trends of cardiovascular medicines consumed by outpatients at JKCI in the fiscal years 2017/2018 to 2021/2022 in defined daily doses per 100 outpatient-days at ATC level 2 classification.

### ATC level C distribution

Across the ATC level 3 subclasses, the cumulative utilisation totalled 360.55 DDD per 100 outpatient days over the study period. The two largest contributors were plain angiotensin II receptor blockers (C09C) and vascular-selective calcium channel blockers (C08C), each exceeding 65 DDD per 100 outpatient days cumulatively (65.46 and 65.62, respectively). ARB + diuretic combinations (C09D) ranked second highest (44.91), followed by centrally acting antiadrenergics (C02A; 38.45), lipid-modifying agents (C10A; 37.58), and high-ceiling diuretics (C03C; 30.65) (Table 2).

**Table 2:**
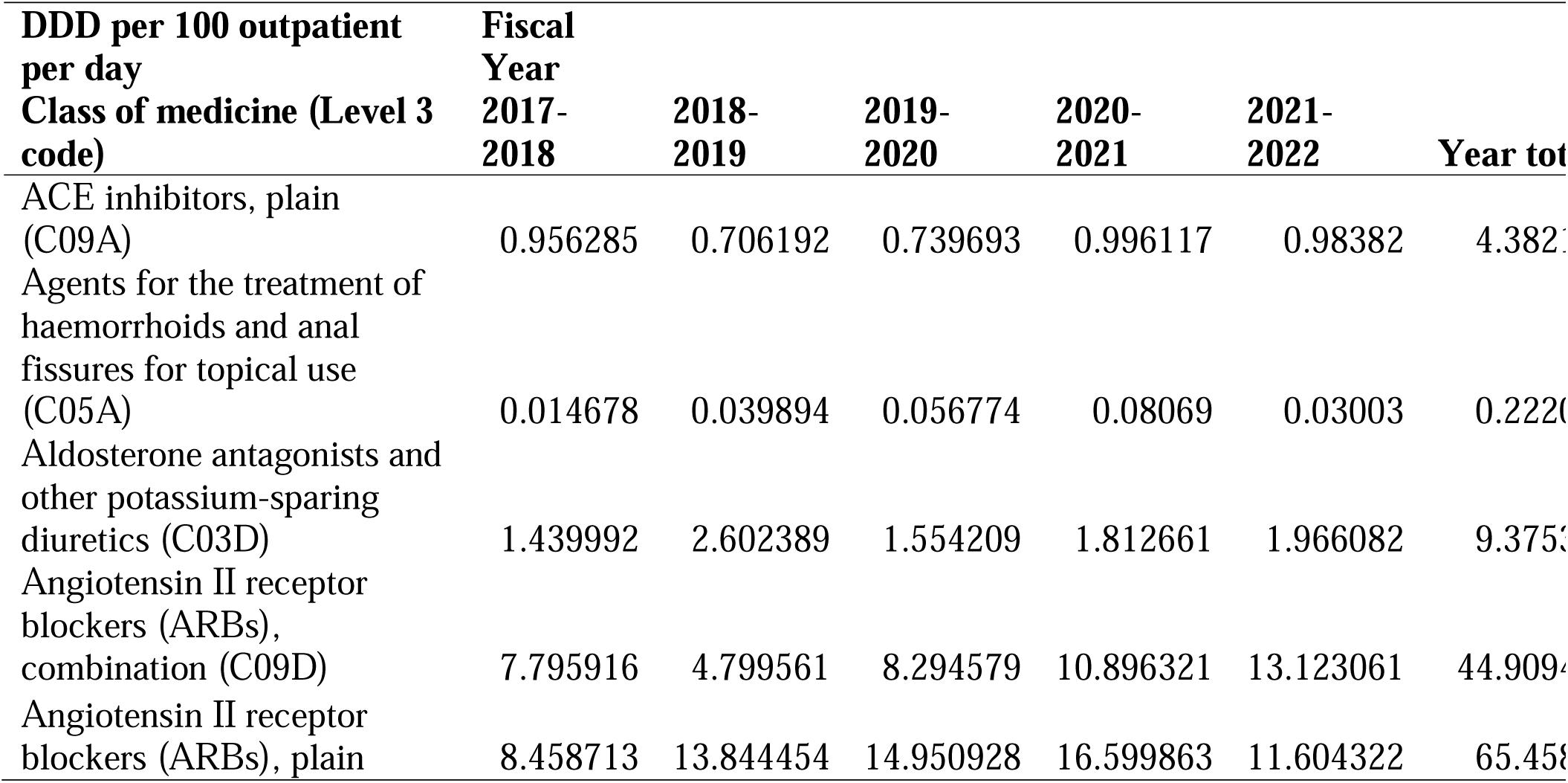
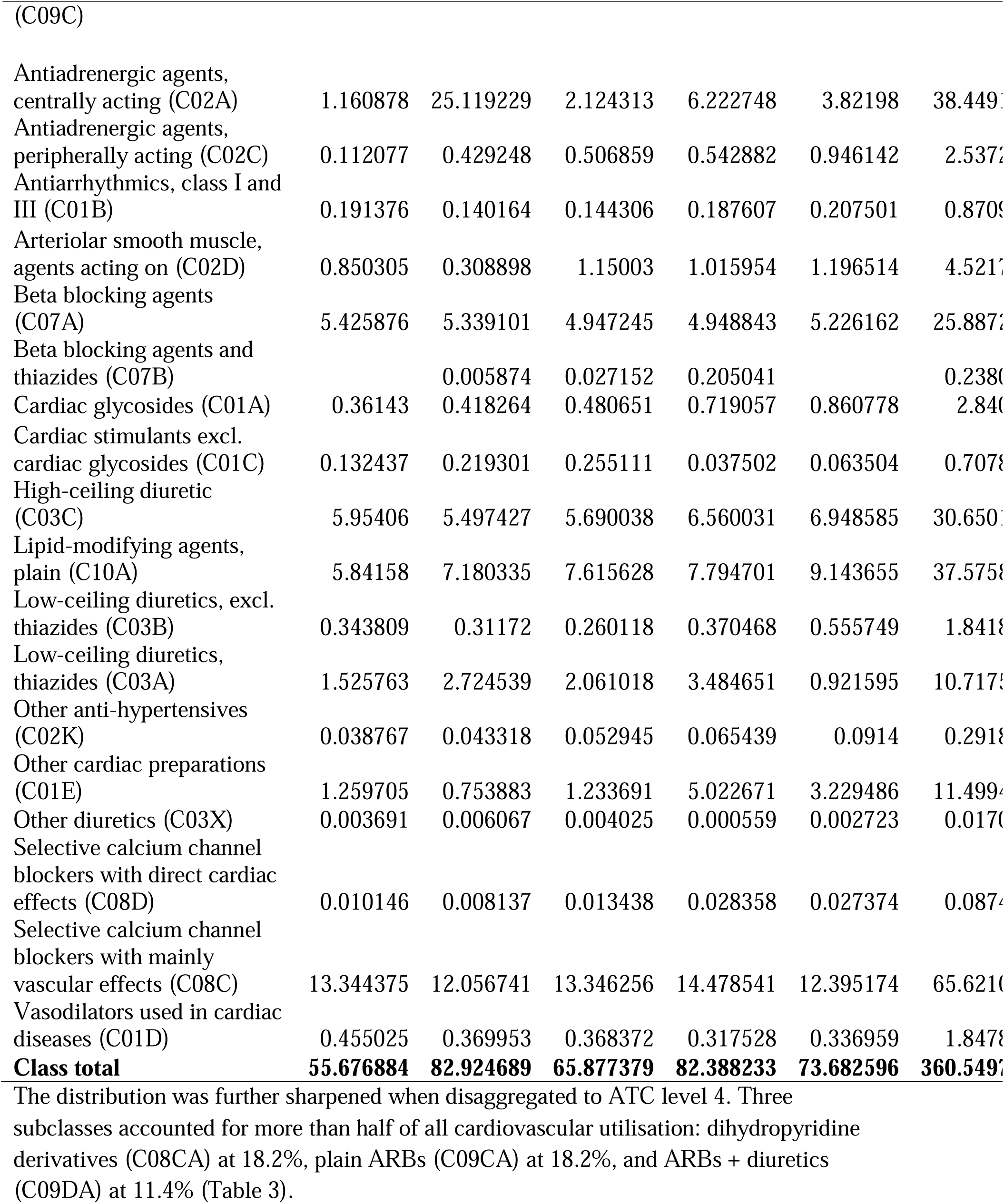
Trends in cardiovascular medicine consumption at JKCI by ATC level 3 category, expressed as DDD per 100 outpatient-days across fiscal years 2017/2018 to 2021/2022 (ATC Level C subclasses).

**Table 3.** Distribution of cardiovascular medicines at JKCI by ATC level 4 category, expressed as cumulative DDD per 100 outpatient-days (fiscal years 2017/2018–2021/2022).

| Rank | Medicine generic name (level 4 code) | DDD per 100 outpatient per day | % | Cumulative % |
| --- | --- | --- | --- | --- |
| 1 | Dihydropyridine derivatives (C08CA) | 65.621086 | 18.2 | 18.2 |
| 2 | Angiotensin II receptor blockers (ARBs), plain (C09CA) | 65.458281 | 18.2 | 36.4 |
| 3 | Angiotensin II receptor blockers (ARBs) and diuretics (C09DA) | 41.256311 | 11.4 | 47.8 |
| 4 | Imidazoline receptor agonists (C02AC) | 38.344958 | 10.6 | 58.4 |
| 5 | HMG CoA reductase inhibitors (C10AA) | 37.5759 | 10.4 | 68.9 |
| 6 | Sulfonamides, plain (C03CA) | 30.650141 | 8.5 | 77.4 |
| 7 | Beta-blocking agents, selective (C07AB) | 21.435612 | 5.9 | 83.3 |
| 8 | Other cardiac preparations (C01EB) | 11.499437 | 3.2 | 86.5 |
| 9 | Thiazides, plain (C03AA) | 10.717566 | 3.0 | 89.5 |
| 10 | Aldosterone antagonists (C03DA) | 9.375333 | 2.6 | 92.1 |
| 11 | Hydrazinophthalazine derivatives (C02DB) | 4.521702 | 1.3 | 93.3 |
| 12 | ACE inhibitors, plain (C09AA) | 4.382107 | 1.2 | 94.5 |
| 13 | Alpha and beta blocking agents (C07AG) | 4.025945 | 1.1 | 95.6 |
| 14 | Angiotensin II receptor blockers (ARBs), and other combinations (C09DX) | 3.653126 | 1.0 | 96.7 |
| 15 | Digitalis glycosides (C01AA) | 2.84018 | 0.8 | 97.5 |
| 16 | Alpha-adrenoreceptor antagonists (C02CA) | 2.537209 | 0.7 | 98.2 |
| 17 | Organic nitrates (C01DA) | 1.847752 | 0.5 | 98.7 |
| 18 | Sulfonamides, plain (C03BA) | 1.841864 | 0.5 | 99.2 |
| 19 | Antiarrhythmics, class III (C01BD) | 0.870917 | 0.2 | 99.4 |
| 20 | Other cardiac stimulants (C01CX) | 0.691528 | 0.2 | 99.6 |
| 21 | Beta-blocking agents, non-selective (C07AA) | 0.42567 | 0.1 | 99.7 |
| 22 | Anti-hypertensive for pulmonary arterial hypertension (C02KX) | 0.291867 | 0.1 | 99.8 |
| 23 | Beta-blocking agents, selective, and thiazides (C07BB) | 0.238067 | 0.1 | 99.9 |
| 24 | Muscle relaxants (C05AE) | 0.222066 | 0.1 | 99.9 |
| 25 | Methyldopa (C02AB) | 0.104189 | 0.0 | 100.0 |
| 26 | Phenylalkylamine derivatives (C08DA) | 0.087453 | 0.0 | 100.0 |
| 27 | Vasopressin antagonists (C03XA) | 0.017065 | 0.0 | 100.0 |
| 28 | Adrenergic and dopaminergic agents (C01CA) | 0.016293 | 0.0 | 100.0 |
| 29 | Other vasodilators used in cardiac diseases (C01DX) | 0.000085 | 0.0 | 100.0 |
| 30 | Antiarrhythmics, class Ic (C01BC) | 0.000037 | 0.0 | 100.0 |
| 31 | Phosphodiesterase inhibitors (C01CE) | 0.000034 | 0.0 | 100.0 |
| **Total** |  | **360.549781** |  |  |

Most other subclasses contributed < 12 DDD per 100 outpatient-days each, with some groups (e.g. topical haemorrhoid preparations C05A and “other diuretics” C03X) contributing < 0.3 cumulatively. These values indicate that relatively few subclasses accounted for the majority of the total cardiovascular utilisation volume at the JKCI.

For cardiovascular medicines at ATC level 5, amlodipine (C08CA01) contributed the highest cumulative utilisation (50.62 DDD per 100 outpatient-days), followed by candesartan (C09CA06) and clonidine (C02AC01) (Table 4). The DU50 segment comprised the top five agents (amlodipine, candesartan, clonidine, furosemide and atorvastatin), which together accounted for approximately 52% of the total cardiovascular utilisation. In contrast, the DU90 segment included the top 18 medicines (cumulative 90.2% of total use). The ranking based on prescription frequency was similar, with amlodipine (C08CA01) being the most frequently dispensed, followed by candesartan (C09CA06), furosemide (C03CA01), spironolactone (C03DA01) and bisoprolol (C07AB07) (Table 4). Distribution of cardiovascular medicines (ATC level 5) dispensed to outpatients at JKCI, ranked by cumulative DDD per 100 outpatient-days (total = 360.55), fiscal years 2017/2018–2021/2022.

**Table 4:**
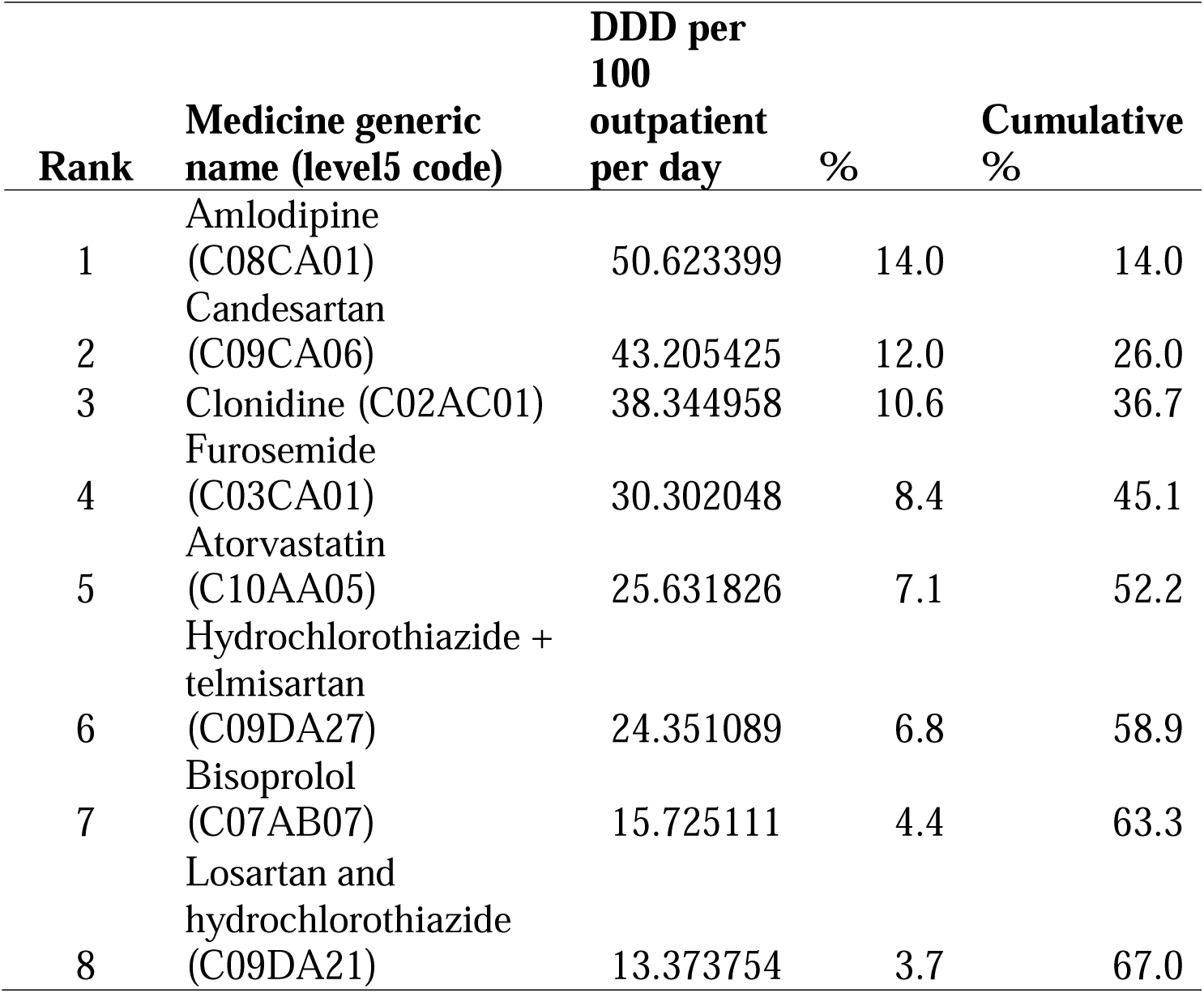
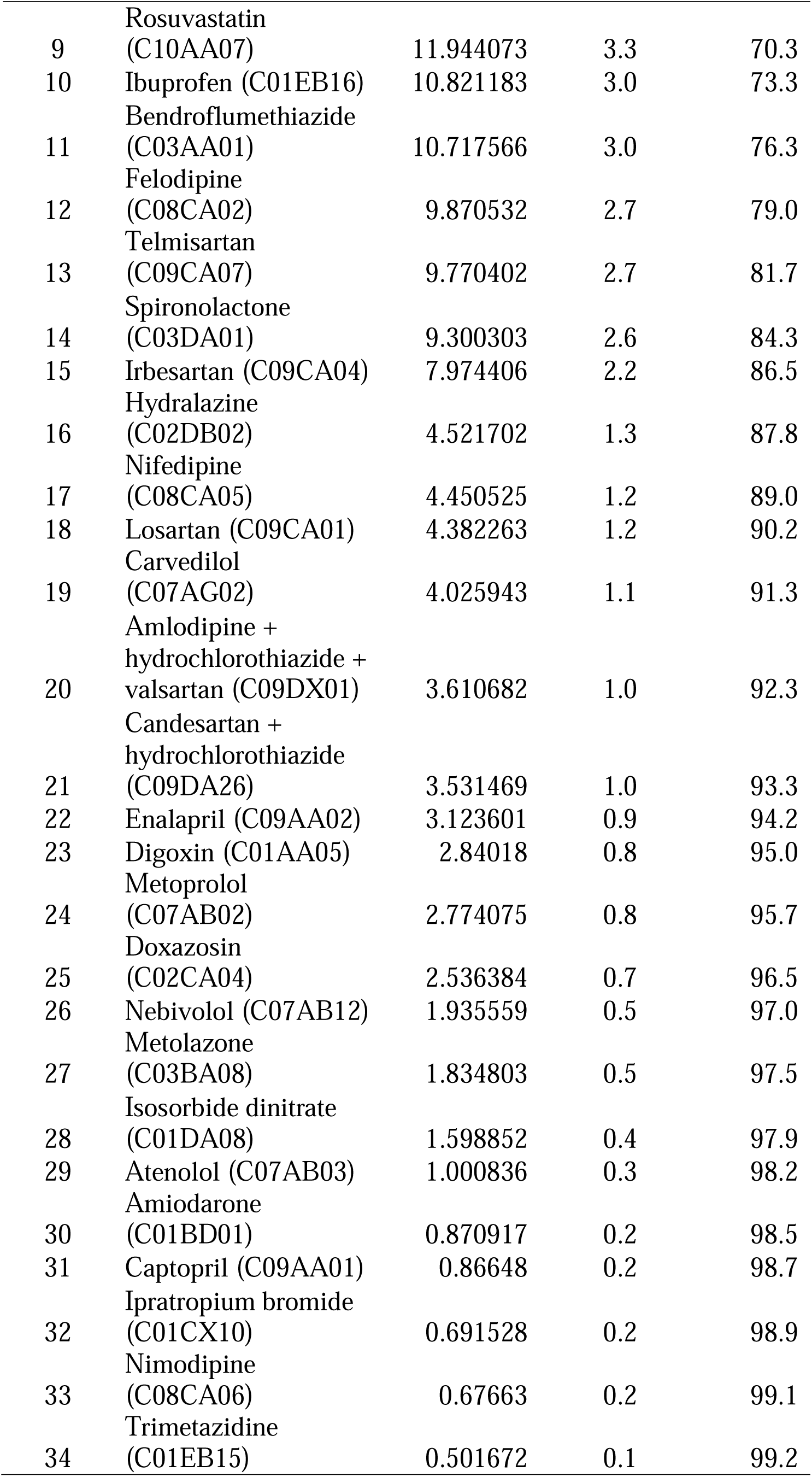
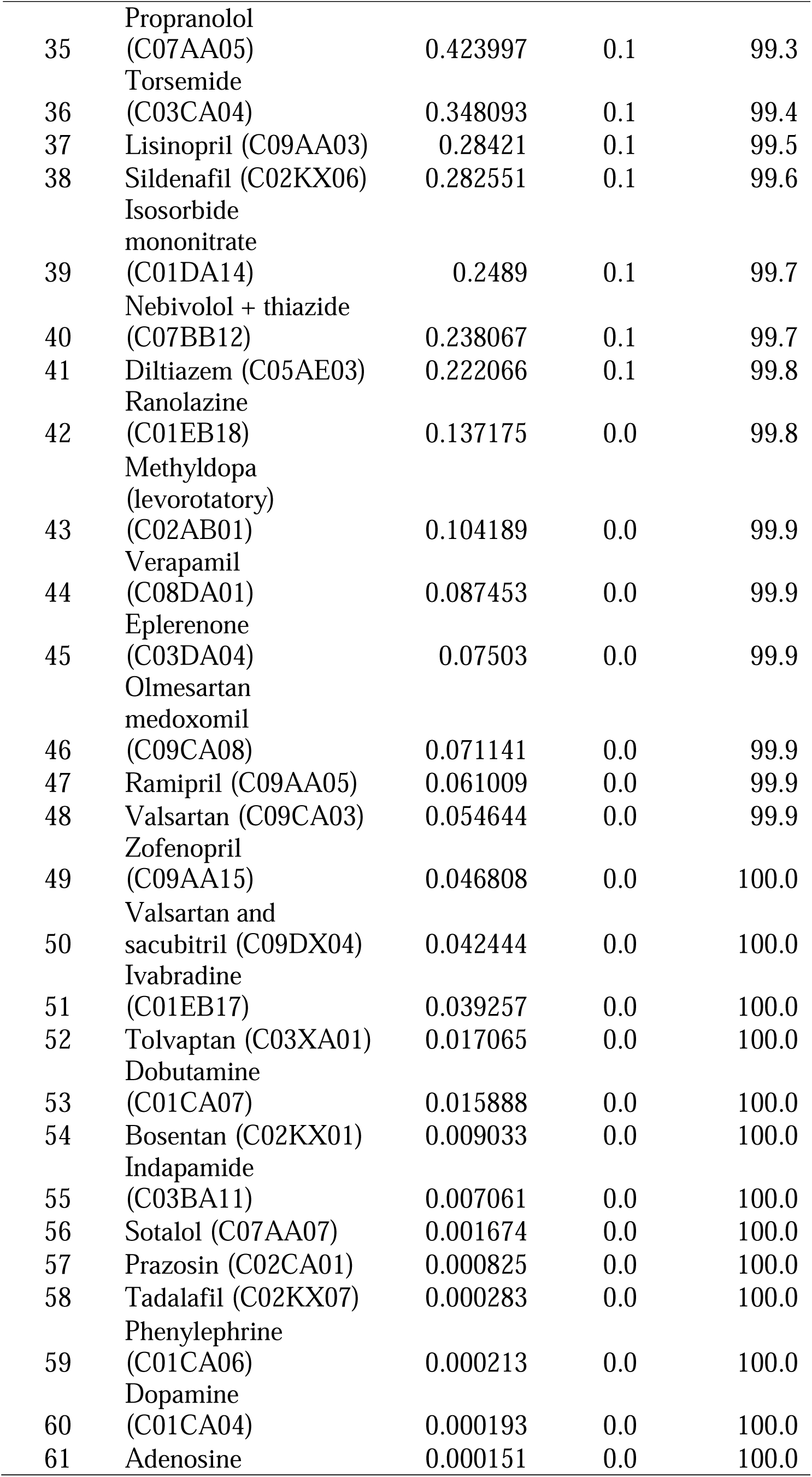
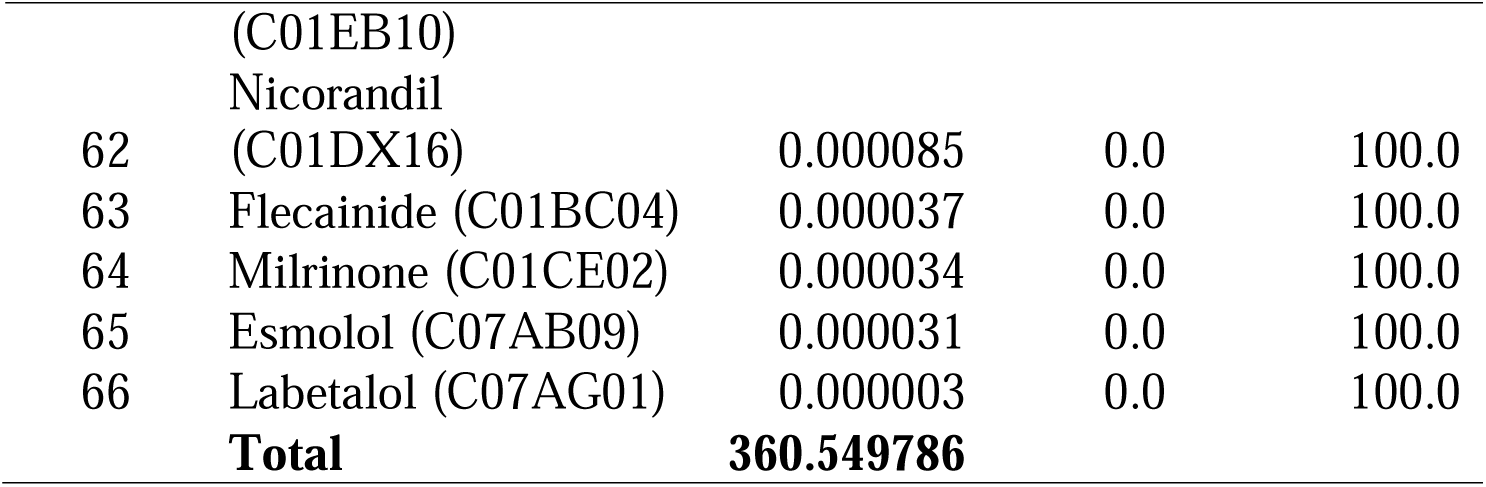
Cardiovascular medicines at the ATC level 5 ranked by cumulative utilisation expressed as DDD per 100 outpatient-days (total = 360.55) over the 2017/2018–2021/2022 period at JKCI. The DU50 boundary occurred for the fifth-ranked substances (amlodipine, candesartan, clonidine, furosemide, and atorvastatin). The DU90 boundary was observed at the 18th-ranked substance.

Monthly utilisation varied over time, with observed monthly values ranging from approximately 3.9 to 6.0 DDD per 100 outpatient days during mid-2017 and from approximately 5.5 to 8.3 DDD per 100 outpatient days during 2021–2022.

The monthly series exhibited pronounced short-term fluctuations and a visible long-range upward trend; the Seasonal–Trend decomposition procedure based on Loess (STL) estimated a trend slope of approximately +0.46 DDD per 100 outpatient days per year. However, there was no statistically significant month-specific seasonal pattern (one-way ANOVA by calendar month, p = 0.923). A single high value was observed in November 2018; however, this represented an isolated month-level anomaly and did not form part of a repeating, seasonal pattern. (Figure 5).

**Figure 5:**
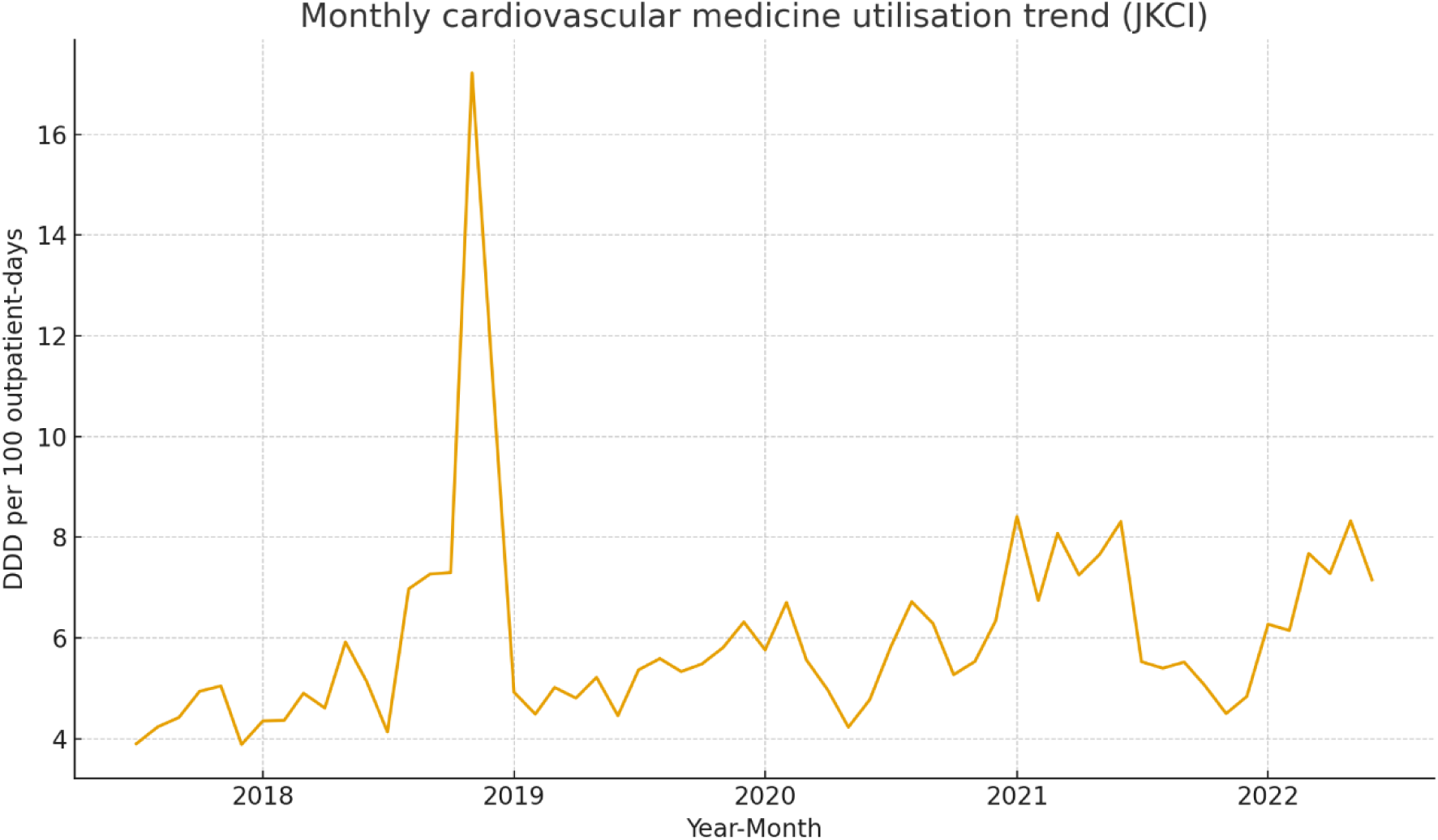
Monthly cardiovascular medicine utilisation (DDD per 100 outpatient-days) at JKCI (July 2017 – June 2022). The Seasonal trend decomposition procedure based on Loess (STL) demonstrated a gradual upward trend over time (∼+0.46 DDD per 100 outpatient days per year), while no statistically significant month-specific seasonal pattern was detected (one-way ANOVA by calendar month p = 0.923).

## 4.0 Discussion

This study provides the first outpatient-based ATC/DDD characterisation of cardiovascular medicine utilisation at a tertiary cardiac centre in Tanzania over five years (2017/2018–2021/2022). Cardiovascular agents consistently formed the dominant therapeutic class at JKCI across all years, consistent with the fact that outpatient pharmacy activity is largely CVD-driven. The annual utilisation intensity (DDD per 100 outpatient days) declined over the study period. In contrast, the composition of ATC subclasses remained relatively stable, suggesting changes in volume rather than structural substitutions of pharmacological groups. The results show consistent annual increases in cardiovascular drug consumption from 2017 to 2022, mirroring trends in other LMICs undergoing an epidemiological transition toward chronic non-communicable diseases [1,4,8].

Within the cardiovascular system (Anatomical Therapeutic Chemical [ATC] level C), utilisation was concentrated within renin–angiotensin system agents (C09), vascular calcium-channel blockers (C08C), high-ceiling diuretics (C03C) and lipid-modifying agents (C10A). Disaggregation to the chemical substance level (ATC level 5) confirmed that amlodipine, candesartan, and clonidine were the principal drivers of cumulative utilisation. DU-segmentation supported this pattern: the DU50 segment comprised only five molecules (amlodipine, candesartan, clonidine, furosemide, and atorvastatin), whereas the DU90 boundary occurred at the 18th-ranked substance, indicating a concentrated therapeutic profile consistent with a hypertension-dominant outpatient case-mix. This aligns with international experience: amlodipine is also the most frequently prescribed anti-hypertensive in several outpatient studies from Ethiopia and India, reflecting the WHO first-line positioning for hypertension and the favourable cost-effectiveness profile of calcium-channel blockers in routine care [6,7,15]. Conversely, the lower utilisation of plain ARBs, such as losartan, has also been reported in LMIC settings, where affordability constraints and formulary positioning influence the uptake of this class [2,9,16].

The high utilisation of cardiovascular medicines among patients aged ≥ 60 years aligns with global demographic patterns showing a disproportionate burden of cardiovascular disease in older adults [10,11]. Comparable age-related utilisation profiles have also been reported in previous inpatient studies from tertiary care settings, reflecting increasing comorbidity and treatment intensity with advancing age [17,18]. The predominance of female patients in this outpatient cohort is consistent with evidence that postmenopausal women experience a marked increase in cardiometabolic risk, including a higher susceptibility to atherothrombotic events, thereby increasing the likelihood of receiving long-term cardiovascular pharmacotherapy [5,10].

The monthly time-series analysis did not identify statistically significant calendar-month seasonal effects (one-way ANOVA, p = 0.923). However, STL demonstrated a gradual, positive underlying trend of approximately +0.46 DDD per 100 outpatient days per year, with a single isolated peak in November 2018. Thus, short-term variation appears irregular rather than cyclical, and medium-term change is better characterised as secular drift rather than seasonality. These findings are consistent with pharmacoepidemiological observations that outpatient maintenance therapy for chronic cardiovascular conditions typically shows weaker seasonality than acute-care medicines (e.g. anti-infectives) [19]. The increasing DDD trends observed for cardiovascular medicines indicate improved detection and management of chronic diseases at JKCI. However, the data also suggest opportunities to strengthen rational prescribing and adherence to the guidelines.

International experience underscores that routine medicine use surveillance contributes to better procurement planning, reduced wastage, and more equitable access to essential medicines [12,13,19]. Our observation of increasing utilisation intensity over time aligns with recent evidence from China, where cardiovascular medicine consumption also increased under policy reform conditions [20]. In Tanzania, institutionalising outpatient DDD monitoring could support both clinical governance and health system stewardship.

The interpretation of these findings should recognise that outpatient data from private hospitals and primary care facilities were unavailable and that national outpatient consumption may therefore be underestimated. Notwithstanding this constraint, three therapeutic groups clearly emerged as stewardship priorities in this tertiary outpatient context: calcium channel blockers, diuretics, and renin–angiotensin system agents. Subsequent studies should evaluate whether changes in DDD utilisation are associated with clinical outcomes, including blood pressure control, hospital readmission, and cardiovascular complication rates, to strengthen evidence-based cardiovascular care in Tanzania.

## Declarations

### Data Availability

The data underlying this study are owned by the Jakaya Kikwete Cardiac Institute (JKCI) and cannot be shared publicly. Researchers may request access directly from the JKCI, subject to institutional and ethical approval. Most of the aggregated data are presented in the manuscript and Supplementary Materials.

### Competing Interests

The authors declare no conflicts of interest.

### Funding

This research did not receive any specific grants from any funding agency in the public, commercial, or not-for-profit sectors.

## Notes

### Competing Interest Statement

The authors have declared no competing interest.

