## Supplementary Table for "Utilisation Patterns of Cardiovascular Medicines Among Outpatients at a Tertiary Cardiac Institute in Tanzania (2017-2022)"

**Supplementary Materials**

Supplementary Table 1: Full distribution of outpatient prescription records for all ATC level 5 medicines utilised at Jakaya Kikwete Cardiac Institute from 2017 to 2022, ranked by frequency of use (N = 1,119,041 prescriptions).

| **Rank** | **Medicine generic name (level 5 ATC code** | **N** | **%** | **Cumulative %** |
| --- | --- | --- | --- | --- |
| 1 | Amlodipine (C08CA01) | 79,357 | 7.1 | 7.1 |
| 2 | Candesartan (C09CA06) | 65,996 | 5.9 | 13.0 |
| 3 | Furosemide (C03CA01) | 64,013 | 5.7 | 18.7 |
| 4 | Spironolactone (C03DA01) | 60,038 | 5.4 | 24.1 |
| 5 | Bisoprolol (C07AB07) | 57,187 | 5.1 | 29.2 |
| 6 | Atorvastatin (C10AA05) | 56,626 | 5.1 | 34.2 |
| 7 | Acetylsalicylic acid (A01AD05) | 54,538 | 4.9 | 39.1 |
| 8 | Hydrochlorothiazide + telmisartan (C09DA27) | 36,208 | 3.2 | 42.4 |
| 9 | Multivitamins + other combinations (A11AB50) | 31,280 | 2.8 | 45.1 |
| 10 | Clopidogrel (B01AC04) | 30,508 | 2.7 | 47.9 |
| 11 | Losartan and hydrochlorothiazide (C09DA21) | 28,730 | 2.6 | 50.4 |
| 12 | Meloxicam (M01AC06) | 27,814 | 2.5 | 52.9 |
| 13 | Carvedilol (C07AG02) | 27,309 | 2.4 | 55.4 |
| 14 | Pregabalin (N03AX16) | 24,688 | 2.2 | 57.6 |
| 15 | Esomeprazole (A02BC05) | 19,552 | 1.7 | 59.3 |
| 16 | Metoprolol (C07AB02) | 18,692 | 1.7 | 61.0 |
| 17 | Hydralazine (C02DB02) | 15,214 | 1.4 | 62.4 |
| 18 | Rosuvastatin (C10AA07) | 14,639 | 1.3 | 63.7 |
| 19 | Glimepiride (A10BB12) | 14,454 | 1.3 | 65.0 |
| 20 | Montelukast (R03DC03) | 14,334 | 1.3 | 66.2 |
| 21 | Aluminium hydroxide + magnesium hydroxide(A02AD10) | 14,329 | 1.3 | 67.5 |
| 22 | Enalapril (C09AA02) | 14,203 | 1.3 | 68.8 |
| 23 | Irbesartan (C09CA04) | 14,167 | 1.3 | 70.0 |
| 24 | Felodipine (C08CA02) | 14,135 | 1.3 | 71.3 |
| 25 | Telmisartan (C09CA07) | 13,938 | 1.2 | 72.6 |
| 26 | Isosorbide dinitrate (C01DA08) | 13,105 | 1.2 | 73.7 |
| 27 | Pantoprazole (A02BC02) | 12,658 | 1.1 | 74.9 |
| 28 | Rabeprazole (A02BC04) | 12,234 | 1.1 | 76.0 |
| 29 | Bendroflumethiazide (C03AA01) | 11,815 | 1.1 | 77.0 |
| 30 | Losartan (C09CA01) | 10,069 | 0.9 | 77.9 |
| 31 | Nifedipine (C08CA05) | 9,264 | 0.8 | 78.7 |
| 32 | Warfarin (B01AA03) | 9,001 | 0.8 | 79.5 |
| 33 | Multivitamins and calcium (A11AA02) | 8,814 | 0.8 | 80.3 |
| 34 | Digoxin (C01AA05) | 8,365 | 0.7 | 81.1 |
| 35 | Febuxostat (M04AA03) | 8,087 | 0.7 | 81.8 |
| 36 | Amlodipine + hydrochlorothiazide + valsartan (C09DX01) | 7,981 | 0.7 | 82.5 |
| 37 | Paracetamol (N02BE01) | 7,720 | 0.7 | 83.2 |
| 38 | Amoxicillin (J01CA04) | 7,545 | 0.7 | 83.9 |
| 39 | Candesartan + hydrochlorothiazide (C09DA26) | 7,409 | 0.7 | 84.5 |
| 40 | Metformin (A10BA02) | 7,111 | 0.6 | 85.2 |
| 41 | Gabapentin (N03AX12) | 5,693 | 0.5 | 85.7 |
| 42 | Loratadine (R06AX13) | 5,601 | 0.5 | 86.2 |
| 43 | Guaifenesin (R05CA03) | 5,514 | 0.5 | 86.7 |
| 44 | Doxazosin (C02CA04) | 5,469 | 0.5 | 87.2 |
| 45 | Omeprazole (A02BC01) | 5,401 | 0.5 | 87.6 |
| 46 | Glucosamine (M01AX05) | 5,074 | 0.5 | 88.1 |
| 47 | Captopril (C09AA01) | 4,804 | 0.4 | 88.5 |
| 48 | Amitriptyline (N06AA09) | 4,546 | 0.4 | 88.9 |
| 49 | Nebivolol (C07AB12) | 4,090 | 0.4 | 89.3 |
| 50 | Cetirizin (R06AE07) | 4,071 | 0.4 | 89.7 |
| 51 | Metolazone (C03BA08) | 3,902 | 0.3 | 90.0 |
| 52 | Rivaroxaban (B01AF01) | 3,846 | 0.3 | 90.4 |
| 53 | Propranolol (C07AA05) | 3,819 | 0.3 | 90.7 |
| 54 | Desloratadine (R06AX27) | 3,621 | 0.3 | 91.0 |
| 55 | Atenolol (C07AB03) | 3,489 | 0.3 | 91.3 |
| 56 | Albendazole (P02CA03) | 3,469 | 0.3 | 91.6 |
| 57 | Prednisolone (A01AC04) | 3,436 | 0.3 | 92.0 |
| 58 | Vildagliptin (A10BH02) | 3,280 | 0.3 | 92.2 |
| 59 | Salbutamol (R03AC02) | 3,121 | 0.3 | 92.5 |
| 60 | Ibuprofen (C01EB16) | 2,977 | 0.3 | 92.8 |
| 61 | Betahistine (N07CA01) | 2,781 | 0.2 | 93.0 |
| 62 | Ferrous sulphate (B03AD03) | 2,616 | 0.2 | 93.3 |
| 63 | Glibenclamid + metformin (A10BD31) | 2,613 | 0.2 | 93.5 |
| 64 | Allopurinol (M04AA01) | 2,554 | 0.2 | 93.7 |
| 65 | Azithromycin (J01FA10) | 2,326 | 0.2 | 93.9 |
| 66 | Glibenclamide (A10BB01) | 2,182 | 0.2 | 94.1 |
| 67 | Menthol (R01AX23) | 2,058 | 0.2 | 94.3 |
| 68 | Tamsulosin (G04CA02) | 2,029 | 0.2 | 94.5 |
| 69 | Sodium chloride (A12CA01) | 2,016 | 0.2 | 94.7 |
| 70 | Phenoxymethylpenicillin-benzathine (J01CE10) | 1,955 | 0.2 | 94.9 |
| 71 | Amiodarone (C01BD01) | 1,826 | 0.2 | 95.0 |
| 72 | Ferrous fumarate (B03AA02) | 1,814 | 0.2 | 95.2 |
| 73 | Budesonide (A07EA06) | 1,750 | 0.2 | 95.3 |
| 74 | Flucloxacillin (J01CF05) | 1,747 | 0.2 | 95.5 |
| 75 | Carbamazepine (N03AF01) | 1,628 | 0.1 | 95.6 |
| 76 | Heparin (B01AB01) | 1,529 | 0.1 | 95.8 |
| 77 | Folic acid (B03BB01) | 1,522 | 0.1 | 95.9 |
| 78 | Isosorbide mononitrate (C01DA14) | 1,506 | 0.1 | 96.0 |
| 79 | Sildenafil (C02KX06) | 1,442 | 0.1 | 96.2 |
| 80 | Insulin aspart (A10AB05) | 1,404 | 0.1 | 96.3 |
| 81 | Fluconazole (D01AC15) | 1,329 | 0.1 | 96.4 |
| 82 | Amoxicillin + clavulanic (J01CR02) | 1,317 | 0.1 | 96.5 |
| 83 | Multivitamins and other minerals, incl. combinations (A11AA03) | 1,316 | 0.1 | 96.7 |
| 84 | Torsemide (C03CA04) | 1,286 | 0.1 | 96.8 |
| 85 | Fluticasone + salmeterol (R03AK06) | 1,183 | 0.1 | 96.9 |
| 86 | Ciprofloxacin (J01MA02) | 1,182 | 0.1 | 97.0 |
| 87 | Finasteride (D11AX10) | 1,133 | 0.1 | 97.1 |
| 88 | Zinc sulfate (A12CB01) | 1,107 | 0.1 | 97.2 |
| 89 | Ampicloxacillin (J01CA51) | 1,085 | 0.1 | 97.3 |
| 90 | Asprin + clopidogrel (B01AC34) | 1,066 | 0.1 | 97.4 |
| 91 | Tramadol (N02AX02) | 1,016 | 0.1 | 97.5 |
| 92 | Cefixime (J01DD08) | 972 | 0.1 | 97.6 |
| 93 | Ceftriaxone (J01DD04) | 968 | 0.1 | 97.6 |
| 94 | Sitagliptin (A10BH01) | 953 | 0.1 | 97.7 |
| 95 | Carbimazole (H03BB01) | 952 | 0.1 | 97.8 |
| 96 | Fluticasone (D07AC17) | 934 | 0.1 | 97.9 |
| 97 | Terbinafine (D01AE15) | 913 | 0.1 | 98.0 |
| 98 | Verapamil (C08DA01) | 903 | 0.1 | 98.1 |
| 99 | Metronidazole (J01XD01) | 878 | 0.1 | 98.1 |
| 100 | Ranolazine (C01EB18) | 837 | 0.1 | 98.2 |
| 101 | Levothyroxine (H03AA01) | 822 | 0.1 | 98.3 |
| 102 | Clarithromycin (J01FA09) | 778 | 0.1 | 98.4 |
| 103 | Trimetazidine (C01EB15) | 736 | 0.1 | 98.4 |
| 104 | Lisinopril (C09AA03) | 686 | 0.1 | 98.5 |
| 105 | Cefpodoxime (J01DD13) | 661 | 0.1 | 98.5 |
| 106 | Lactulose (A06AD11) | 614 | 0.1 | 98.6 |
| 107 | Ketoprofen (M01AE03) | 586 | 0.1 | 98.6 |
| 108 | Domperidone (A03FA03) | 554 | 0.0 | 98.7 |
| 109 | Mebendazole (P02CA01) | 552 | 0.0 | 98.7 |
| 110 | Levodopa (N04BA01) | 551 | 0.0 | 98.8 |
| 111 | Pioglitazone (A10BG03) | 549 | 0.0 | 98.8 |
| 112 | Bisacodyl (A06AB02) | 540 | 0.0 | 98.9 |
| 113 | Ketoconazole (D01AC08) | 540 | 0.0 | 98.9 |
| 114 | Ornidazole (G01AF06) | 449 | 0.0 | 99.0 |
| 115 | Artemisinin piperaquine(P01BF07) | 436 | 0.0 | 99.0 |
| 116 | Artemether + lumefantrine (P01BF01) | 394 | 0.0 | 99.1 |
| 117 | Miconazole (J02AB01) | 377 | 0.0 | 99.1 |
| 118 | Methyldopa (levorotatory) (C02AB01) | 370 | 0.0 | 99.1 |
| 119 | Diltiazem (C05AE03) | 366 | 0.0 | 99.2 |
| 120 | Cefalexin (J01DB01) | 338 | 0.0 | 99.2 |
| 121 | Potassium chloride (A12BA01) | 332 | 0.0 | 99.2 |
| 122 | Aciclovir (D06BB03) | 326 | 0.0 | 99.2 |
| 123 | Eplerenone (C03DA04) | 320 | 0.0 | 99.3 |
| 124 | Tolvaptan (C03XA01) | 320 | 0.0 | 99.3 |
| 125 | Bromazepam (N05BA08) | 285 | 0.0 | 99.3 |
| 126 | Piperacillin + tazobactam (J01CR05) | 282 | 0.0 | 99.3 |
| 127 | Tinidazole (J01XD02) | 264 | 0.0 | 99.4 |
| 128 | Artemether (P01BE02) | 260 | 0.0 | 99.4 |
| 129 | Diclofenac (D11AX18) | 251 | 0.0 | 99.4 |
| 130 | Haloperidol (N05AD01) | 234 | 0.0 | 99.4 |
| 131 | Phenobarbital (N03AA02) | 223 | 0.0 | 99.5 |
| 132 | Clonidine (C02AC01) | 222 | 0.0 | 99.5 |
| 133 | Saccharated iron oxide (B03AB02) | 209 | 0.0 | 99.5 |
| 134 | Meropenem (J01DH02) | 206 | 0.0 | 99.5 |
| 135 | Ivabradine (C01EB17) | 203 | 0.0 | 99.5 |
| 136 | Baclofen (M03BX01) | 187 | 0.0 | 99.5 |
| 137 | Gentamicin (J01GB03) | 186 | 0.0 | 99.6 |
| 138 | Nebivolol + thiazide (C07BB12) | 178 | 0.0 | 99.6 |
| 139 | Amoxicillin + metronidazole + omeprazole (A02BD01) | 167 | 0.0 | 99.6 |
| 140 | Vancomycin (J01XA01) | 165 | 0.0 | 99.6 |
| 141 | Metoclopramide (A03FA01) | 155 | 0.0 | 99.6 |
| 142 | Bosentan (C02KX01) | 154 | 0.0 | 99.6 |
| 143 | Hydrocortisone (A01AC03) | 145 | 0.0 | 99.7 |
| 144 | Dapagliflozin (A10BK01) | 138 | 0.0 | 99.7 |
| 145 | Ipratropium bromide (C01CX10) | 137 | 0.0 | 99.7 |
| 146 | Sulfamethoxazole + trimethoprim (J01EE01) | 136 | 0.0 | 99.7 |
| 147 | Artemisinin (P01BE01) | 132 | 0.0 | 99.7 |
| 148 | Adrenaline (A01AD06) | 125 | 0.0 | 99.7 |
| 149 | Thiamine (N07XB52) | 123 | 0.0 | 99.7 |
| 150 | Ipratropium bromide + salbutamol (R03AL02) | 120 | 0.0 | 99.7 |
| 151 | Valsartan and sacubitril (C09DX04) | 112 | 0.0 | 99.7 |
| 152 | Zofenopril (C09AA15) | 112 | 0.0 | 99.8 |
| 153 | Aceclofenac (M01AB16) | 110 | 0.0 | 99.8 |
| 154 | Ferrous sulphate + folic acid (B03BB51) | 107 | 0.0 | 99.8 |
| 155 | Insulin (human) (A10AB01) | 104 | 0.0 | 99.8 |
| 156 | Loperamide (A07DA03) | 100 | 0.0 | 99.8 |
| 157 | Ondansetron (A04AA01) | 94 | 0.0 | 99.8 |
| 158 | Ascorbic acid (G01AD03) | 87 | 0.0 | 99.8 |
| 159 | Olmesartan medoxomil (C09CA08) | 86 | 0.0 | 99.8 |
| 160 | Povidone iodine (S01AX18) | 84 | 0.0 | 99.8 |
| 161 | Dexamethasone (A01AC02) | 75 | 0.0 | 99.8 |
| 162 | Diphenhydramine (A04AB05) | 73 | 0.0 | 99.8 |
| 163 | Doxycycline (J01AA02) | 73 | 0.0 | 99.8 |
| 164 | Itraconazole (J02AC02) | 69 | 0.0 | 99.8 |
| 165 | Morphine (N02AA01) | 69 | 0.0 | 99.9 |
| 166 | Diclofenac + paracetamol + serratiopeptidase (M01AB55) | 68 | 0.0 | 99.9 |
| 167 | Doxorubicin (L01DB01) | 68 | 0.0 | 99.9 |
| 168 | Ampicillin (J01CA01) | 66 | 0.0 | 99.9 |
| 169 | Ramipril (C09AA05) | 66 | 0.0 | 99.9 |
| 170 | Dobutamine (C01CA07) | 59 | 0.0 | 99.9 |
| 171 | Artesunate (P01BE03) | 53 | 0.0 | 99.9 |
| 172 | Tocopherol (vitamin E) (A11HA03) | 50 | 0.0 | 99.9 |
| 173 | Mannitol (A06AD16) | 49 | 0.0 | 99.9 |
| 174 | Atropine (A03BA01) | 45 | 0.0 | 99.9 |
| 175 | Albumin (B05AA01) | 44 | 0.0 | 99.9 |
| 176 | Calcium gluconate (A12AA03) | 44 | 0.0 | 99.9 |
| 177 | Sodium lactate(B05XA08) | 42 | 0.0 | 99.9 |
| 178 | Diazepam (N05BA01) | 36 | 0.0 | 99.9 |
| 179 | Erythromycin (J01FA01) | 36 | 0.0 | 99.9 |
| 180 | Glycerol (A06AG04) | 36 | 0.0 | 99.9 |
| 181 | Glyceryl trinitrate (A05AX08) | 35 | 0.0 | 99.9 |
| 182 | Lorazepam (N05BA06) | 34 | 0.0 | 99.9 |
| 183 | Streptokinase (B01AD01) | 34 | 0.0 | 99.9 |
| 184 | Tranexamic acid (B02AA02) | 34 | 0.0 | 99.9 |
| 185 | Midazolam (N03AE02) | 33 | 0.0 | 99.9 |
| 186 | Sodium bicarbonate (B05CB04) | 33 | 0.0 | 99.9 |
| 187 | Cefuroxime (J01DC02) | 29 | 0.0 | 99.9 |
| 188 | Dextrose (B05AA05) | 27 | 0.0 | 99.9 |
| 189 | Trihexyphenidyl (N04AA01) | 27 | 0.0 | 99.9 |
| 190 | Valsartan (C09CA03) | 27 | 0.0 | 100.0 |
| 191 | Indapamide (C03BA11) | 26 | 0.0 | 100.0 |
| 192 | Phenytoin (N03AB02) | 26 | 0.0 | 100.0 |
| 193 | Hyoscine butylbromide (A03BB01) | 25 | 0.0 | 100.0 |
| 194 | Sodium citrate (B05CB02) | 23 | 0.0 | 100.0 |
| 195 | Triamcinolone (A01AC01) | 23 | 0.0 | 100.0 |
| 196 | Betamethasone (A07EA04) | 22 | 0.0 | 100.0 |
| 197 | Griseofulvin (D01AA08) | 22 | 0.0 | 100.0 |
| 198 | Paracetamol, combination (N02CX63) | 22 | 0.0 | 100.0 |
| 199 | Acetazolamide (S01EC01) | 21 | 0.0 | 100.0 |
| 200 | Calcium carbonate (A02AC01) | 21 | 0.0 | 100.0 |
| 201 | Phytomenadione (B02BA01) | 19 | 0.0 | 100.0 |
| 202 | Phenoxymethylpenicillin (J01CE02) | 18 | 0.0 | 100.0 |
| 203 | Nystatin (A01AB33) | 16 | 0.0 | 100.0 |
| 204 | Ranitidine (A02BA02) | 15 | 0.0 | 100.0 |
| 205 | Levetiracetam (N03AX14) | 14 | 0.0 | 100.0 |
| 206 | Adenosine (C01EB10) | 12 | 0.0 | 100.0 |
| 207 | Ceftriaxone and beta-lactamase inhibitor (J01DD63) | 12 | 0.0 | 100.0 |
| 208 | Tadalafil (C02KX07) | 11 | 0.0 | 100.0 |
| 209 | Bromocriptine (N04BC01) | 10 | 0.0 | 100.0 |
| 210 | Colchicine (M04AC01) | 10 | 0.0 | 100.0 |
| 211 | Nimodipine (C08CA06) | 10 | 0.0 | 100.0 |
| 212 | Ivermectin (D11AX22) | 9 | 0.0 | 100.0 |
| 213 | Beclomethasone + salbutamol (R03BA01) | 8 | 0.0 | 100.0 |
| 214 | Nicotine (N07BA01) | 8 | 0.0 | 100.0 |
| 215 | Promethazine (D04AA10) | 8 | 0.0 | 100.0 |
| 216 | Valproic acid (N03AG01) | 8 | 0.0 | 100.0 |
| 217 | Lidocaine (A01AE01) | 7 | 0.0 | 100.0 |
| 218 | Paracetamol + tramadol (N02AJ13) | 7 | 0.0 | 100.0 |
| 219 | Secnidazole (P01AB07) | 7 | 0.0 | 100.0 |
| 220 | Clotrimazole, combination (D01AC51) | 6 | 0.0 | 100.0 |
| 221 | Gliclazide (A10BB09) | 6 | 0.0 | 100.0 |
| 222 | Magnesium sulfate (A06AD04) | 6 | 0.0 | 100.0 |
| 223 | Metformin + sitagliptin (A10BD07) | 5 | 0.0 | 100.0 |
| 224 | Minerals (R01AX21) | 5 | 0.0 | 100.0 |
| 225 | Norethisterone (G03AC01) | 5 | 0.0 | 100.0 |
| 226 | Aminophylline (R03DA05) | 4 | 0.0 | 100.0 |
| 227 | Cinnarizine (N06DX12) | 4 | 0.0 | 100.0 |
| 228 | Dopamine (C01CA04) | 4 | 0.0 | 100.0 |
| 229 | Fluoxetine (N06AB03) | 4 | 0.0 | 100.0 |
| 230 | Hydrocodone + paracetamol (N02AJ01) | 4 | 0.0 | 100.0 |
| 231 | Milrinone (C01CE02) | 4 | 0.0 | 100.0 |
| 232 | Esmolol (C07AB09) | 3 | 0.0 | 100.0 |
| 233 | Glucosamine, combination (M01AX55) | 3 | 0.0 | 100.0 |
| 234 | Hydroxycarbamide (L01XX05) | 3 | 0.0 | 100.0 |
| 235 | Octreotide (H01CB02) | 3 | 0.0 | 100.0 |
| 236 | Risperidone (N05AX08) | 3 | 0.0 | 100.0 |
| 237 | Amikacin (J01GB06) | 2 | 0.0 | 100.0 |
| 238 | Cefepime (J01DE01) | 2 | 0.0 | 100.0 |
| 239 | Dexchlorpheniramine, combination (R06AB52) | 2 | 0.0 | 100.0 |
| 240 | Methylprednisolone (D07AA01) | 2 | 0.0 | 100.0 |
| 241 | Nicorandil (C01DX16) | 2 | 0.0 | 100.0 |
| 242 | Prazosin (C02CA01) | 2 | 0.0 | 100.0 |
| 243 | Sevoflurane (N01AB08) | 2 | 0.0 | 100.0 |
| 244 | Sotalol (C07AA07) | 2 | 0.0 | 100.0 |
| 245 | Bleomycin (L01DC01) | 1 | 0.0 | 100.0 |
| 246 | Cefoperazone + salbactam (J01DD62) | 1 | 0.0 | 100.0 |
| 247 | Cilostazol (B01AC23) | 1 | 0.0 | 100.0 |
| 248 | Citicoline (N06BX06) | 1 | 0.0 | 100.0 |
| 249 | Clindamycin (J01FF01) | 1 | 0.0 | 100.0 |
| 250 | Etomidate (N01AX07) | 1 | 0.0 | 100.0 |
| 251 | Flecainide (C01BC04) | 1 | 0.0 | 100.0 |
| 252 | Formoterol and budesonide (R03AK07) | 1 | 0.0 | 100.0 |
| 253 | Glycopyrolate (D11AA01) | 1 | 0.0 | 100.0 |
| 254 | Labetalol (C07AG01) | 1 | 0.0 | 100.0 |
| 255 | Lacosamide (N03AX18) | 1 | 0.0 | 100.0 |
| 256 | Methylcellulose (A06AC06) | 1 | 0.0 | 100.0 |
| 257 | Nalidixic acid (J01MB02) | 1 | 0.0 | 100.0 |
| 258 | Phenylephrine (C01CA06) | 1 | 0.0 | 100.0 |
| 259 | Propofol (N01AX10) | 1 | 0.0 | 100.0 |
| 260 | Protamine (V03AB14) | 1 | 0.0 | 100.0 |
| 261 | Pseudoephedrine and triprolidin (R01BA59) | 1 | 0.0 | 100.0 |
| 262 | Tenecteplase (B01AD11) | 1 | 0.0 | 100.0 |
| 263 | Ticagrelor (B01AC24) | 1 | 0.0 | 100.0 |
| 264 | Tizanidine (M03BX02) | 1 | 0.0 | 100.0 |
| 265 | Undecylenic acid (D01AE04) | 1 | 0.0 | 100.0 |
|  | **Total** | **1,119,041** | **100.0** |  |
